# Women’s mortality following pregnancy affected by vaginal bleeding due to threatened miscarriage: a Danish cohort study

**DOI:** 10.1101/2022.11.25.22282740

**Authors:** Elena Dudukina, Erzsébet Horváth-Puhó, Henrik Toft Sørensen, Vera Ehrenstein

**Affiliations:** Department of Clinical Epidemiology, Aarhus University and Aarhus University Hospital, Aarhus, Denmark

**Keywords:** cohort study, pregnancy, vaginal bleeding, termination, miscarriage, mortality

## Abstract

**Background:** Women with only pregnancy terminations or only miscarriages have an increased mortality risk. We investigated the association between vaginal bleeding (VB) in pregnancy ending in childbirth and women’s mortality.

**Methods:** We conducted a cohort study in Denmark, which included 1,354,181 women and their 3,162,317 pregnancies (1979-2017) followed through 2018. We ascertained 70,835 VB-affected pregnancies and comparators: 2,236,359 VB-unaffected pregnancies ending in childbirth; 589,697 terminations; and 265,940 miscarriages. We computed all-cause and cause-specific mortality rates per 10,000 person-years (PY) and hazard ratios (HRs) with 95% confidence intervals (CIs) using Cox proportional hazards regression adjusted for age, calendar year, preexisting conditions, and socioeconomic factors.

**Results:** There were 2,320 deaths from any cause among women following VB-affected pregnancy (mortality rate: 15.2, 95% CI: 14.6-15.9 per 10,000 PY); 55,030 deaths following VB-unaffected pregnancy (12.7, 12.6-1.28); 27,500 deaths following a termination (21.9, 21.6-22.1), and 10,865 deaths following a miscarriage (19.2, 18.8-19.6).

For comparison of VB-affected vs VB-unaffected pregnancies, associations with all-cause (HR: 1.14, 95% CI: 1.09-1.19), natural-causes (HR: 1.15, 95% CI: 1.09-1.22) and non-natural causes (HR: 1.27, 95% CI: 1.08-1.48) mortality attenuated in a sensitivity analysis of pregnancies recorded in 1994-2017 (HR: 1.00, 95% CI: 0.90-1.12, HR: 0.98, 95% CI: 0.85-1.14, and HR: 1.04, 95% CI: 0.71-1.51, respectively). Contrasts with remaining comparators did not suggest increased risks of all-cause, natural, or non-natural mortality causes.

**Conclusions:** We found no evidence of an increased risk of mortality in women following VB-affected vs VB-unaffected pregnancy, termination, or miscarriage.

**Key Messages:**

- Previous studies mostly focused on short-term outcomes of the newborns and mothers following the vaginal bleeding-affected pregnancy. This study investigated the association between vaginal bleeding in pregnancy ending in childbirth and women’s mortality.
- This registry-based cohort study found no evidence of an increased risk of all-cause or cause-specific mortality from natural or non-natural causes in women following vaginal bleeding-affected pregnancy compared with vaginal bleeding-unaffected pregnancy, termination, or miscarriage

## Introduction

Previous studies showed an association between women’s reproductive health and mortality risk. A Danish study in 1973-2004 reported that a miscarriage or a pregnancy termination was associated with 3-4-fold greater mortality odds compared with a childbirth.^1^ In the same study, women with a history of discordant pregnancy outcomes (childbirths, terminations, and miscarriages) had 1.6-fold greater mortality than women with childbirth only; however, this study did not use time-to-event analysis.^1^ Another Danish study, in 1980-2004, showed that miscarriage of the first pregnancy compared with a childbirth of the first pregnancy was associated with 1.3 and 18.4 additional deaths per 10,000 women at 1 and 10 years of follow-up, respectively.^2^ Recurrent miscarriages are associated with both all-cause and cause-specific mortality in women^3^ possibly via the development of post-traumatic stress disorder, depression, and anxiety.^4^

Threatened miscarriage is a common pregnancy complication^5^ affecting up to 20-30% of clinically recognized pregnancies. Clinically threatened miscarriage manifests as a vaginal bleeding (VB) before 20 gestational weeks in a presence of a viable intrauterine pregnancy.^6^ Potential shared biologic pathways of threatened miscarriage and completed spontaneous miscarriage include low progesterone levels^4,7–13^ and elevated levels of pro-inflammatory cytokines.^14–18^ Although an association between VB and cardiovascular diseases was previously reported,^19^ little is known regarding the association between having a VB-affected pregnancy and women’s mortality.

In this registry-based cohort study, we investigated whether a VB-affected pregnancy ending in childbirth was associated with the woman’s all-cause and cause-specific mortality using three comparators: 1) VB-unaffected pregnancies ending in childbirth, 2) pregnancies ending in a termination, or 3) pregnancies ending in a miscarriage.

## Methods

### Setting, data sources and data linkage

We conducted this study using data linked from health and administrative registries of Denmark, a country with a 2.9 million population of women and universal healthcare access to all its residents. Danish registries collect data prospectively and at an individual level.^20^ The Danish Civil Registration System (CRS) contains the date of birth; vital, civil, and migration status; and a civil registration number, which serves as a unique personal identifier for data linkage and is updated daily.^21^ The data for research are anonymised and stored on the servers of Statistics Denmark.^22^

Information on maternal (age, parity) and pregnancy characteristics (delivery date, gestational age, singleton vs multifetal pregnancy, still-vs live birth) for all deliveries are available in the Danish Medical Birth Registry (MBR) starting from 1973.^23^ Maternal smoking recorded at a first prenatal visit and the body-mass index (BMI) became available in the MBR from 1991 and 2004, respectively.^23^ Data on hospital encounters are available in the Danish National Patient Registry (DNPR), which tracks admission and discharge diagnoses according to the International Classification of Diseases, 10^th^ Revision (ICD-10) starting in 1994 and ICD-8 earlier (1977-1993), procedures, data on outpatient specialist clinics and emergency room visits (from 1995 onwards).^24^ In the DNPR, primary discharge diagnoses reflect the main reason for the visit, while secondary discharge diagnoses give information on other conditions related to the patient visit. The Psychiatric Central Research Register encompasses psychiatric hospital encounters since 1969 and outpatient specialist clinic encounters since 1995 (the latter was available in this study).^25^ The information on medication use including the date of prescription redemption and Anatomical Therapeutic Classification (ATC) code of the prescribed drug is available since 1995 in the Danish National Prescription Registry.^26,27^ Data on the highest completed level of education are captured by the Danish education registers from 1981.^28^ Year-specific personal income at an individual level is available starting in 1980 from the Danish Income Registries.^29^ Danish registers on personal labour market affiliation contain data on employment status from 1980.^30^

### Study population

The study population consisted of pregnancies ending in a delivery, termination, or miscarriage. We ascertained all stillbirths and live births recorded in the MBR between 1979 and 2017. Pregnancies ending in the birth of twins or triplets were analysed as one observation. The exposure to VB due to threatened miscarriage within 20 weeks of gestation ending in delivery was identified using primary and secondary diagnoses recorded in the DNPR^31^at any hospital encounter (Table 1, available as Supplementary data). When several vaginal bleeding episodes were recorded during the first 20 gestational weeks of the same pregnancy, we included the earliest. Pregnancies ending in a childbirth without a record of VB during 20 gestational weeks in the DNPR formed the comparison cohort of VB-unaffected pregnancies. We followed cohorts of VB-affected and VB-unaffected pregnancies from the date of delivery.

**Table 1.** Descriptive characteristics of women with vaginal bleeding (VB)-affected and VB-unaffected pregnancies ending in childbirth, pregnancies ending in a termination or miscarriage, Denmark, 1979-2017.

|  | **VB-affected pregnancy, N=70,835** | **VB-unaffected pregnancy, N=2,236,360** | **Termination, N=589,700** | **Miscarriage, N=265,430** |
| --- | --- | --- | --- | --- |
| **Distinct women** | **N=66,881** | **N=1,198,206** | **N=424,834** | **N=225,103** |
| **Characteristic** | **n (%)<sup>a</sup>** | **n (%)<sup>a</sup>** | **n (%)<sup>a</sup>** | **n (%)<sup>a</sup>** |
| **Calendar year of pregnancy end** |  |  |  |  |
| 1979-1987 | 13,650 (19.0) | 383,605 (17.0) | 142,790 (24.0) | 52,995 (20.0) |
| 1988-1994 | 19,845 (28.0) | 479,240 (21.0) | 120,210 (20.0) | 74,335 (28.0) |
| 1995-2002 | 17,450 (25.0) | 433,245 (19.0) | 122,125 (21.0) | 46,450 (18.0) |
| 2003-2009 | 12,205 (17.0) | 487,340 (22.0) | 117,835 (20.0) | 54,365 (20.0) |
| 2010-2017 | 7,685 (11.0) | 452,930 (20.0) | 86,735 (15.0) | 37,280 (14.0) |
| **Age at the index date, years** |  |  |  |  |
| <20 | 1,460 (2.1) | 49,345 (2.2) | 89,560 (15.0) | 10,370 (3.9) |
| 20-24 | 12,495 (18.0) | 379,360 (17.0) | 140,655 (24.0) | 45,045 (17.0) |
| 25-29 | 25,390 (36.0) | 814,095 (36.0) | 123,745 (21.0) | 79,945 (30.0) |
| 30-34 | 20,975 (30.0) | 680,405 (30.0) | 112,320 (19.0) | 69,845 (26.0) |
| 35-39 | 8,910 (13.0) | 267,135 (12.0) | 85,020 (14.0) | 42,180 (16.0) |
| 40+ | 1,610 (2.3) | 46,015 (2.1) | 38,390 (6.5) | 18,035 (6.8) |
| **Parity at the index date** |  |  |  |  |
| Nulliparous | - | - | 418,295 (71.0) | 226,025 (85.0) |
| Primiparous | 33,450 (47.0) | 1,051,970 (47.0) | 47,150 (8.0) | 16,240 (6.1) |
| Multiparous | 37,385 (53.0) | 1,184,385 (53.0) | 124,255 (21.0) | 23,160 (8.7) |
| **Number of pregnancies at the index date** |  |  |  |  |
| 1 | 22,665 (32.0) | 961,345 (43.0) | 264,630 (45.0) | 103,745 (39.0) |
| 2 | 22,970 (32.0) | 713,415 (32.0) | 115,695 (20.0) | 77,940 (29.0) |
| 3 | 13,380 (19.0) | 337,440 (15.0) | 90,000 (15.0) | 43,415 (16.0) |
| 3+ | 11,820 (17.0) | 224,160 (10.0) | 119,370 (20.0) | 40,325 (15.0) |
| **Civil status** |  |  |  |  |
| Married or in partnership | 41,555 (59.0) | 1,332,090 (60.0) | 201,520 (34.0) | 136,070 (51.0) |
| Not married | 25,000 (35.0) | 759,990 (34.0) | 252,485 (43.0) | 75,630 (28.0) |
| Missing | 4,280 (6.0) | 144,275 (6.5) | 135,690 (23.0) | 53,725 (20.0) |
| **Employment status** |  |  |  |  |
| Employed | 49,910 (70.0) | 1,597,800 (71.0) | 340,660 (58.0) | 183,785 (69.0) |
| Retirement or state pension | 6,455 (9.1) | 170,730 (7.6) | 71,930 (12.0) | 24,785 (9.3) |
| Unemployed | 10,680 (15.0) | 333,615 (15.0) | 128,230 (22.0) | 39,085 (15.0) |
| Missing | 3,785 (5.3) | 134,210 (6.0) | 48,870 (8.3) | 17,770 (6.7) |
| **Highest achieved education** |  |  |  |  |
| Basic education | 21,965 (31.0) | 547,675 (24.0) | 253,135 (43.0) | 78,030 (29.0) |
| High school or similar | 25,590 (36.0) | 840,030 (38.0) | 170,275 (29.0) | 92,675 (35.0) |
| Higher education | 16,575 (23.0) | 625,140 (28.0) | 75,020 (13.0) | 64,185 (24.0) |
| Missing | 6,705 (9.5) | 223,510 (10.0) | 91,265 (15.0) | 30,535 (12.0) |
| **Yearly income, quartiles** |  |  |  |  |
| Q1 | 16,030 (23.0) | 440,100 (20.0) | 219,415 (37.0) | 59,630 (22.0) |
| Q2 | 16,890 (24.0) | 523,855 (23.0) | 130,905 (22.0) | 61,850 (23.0) |
| Q3 | 16,655 (24.0) | 566,255 (25.0) | 90,490 (15.0) | 60,465 (23.0) |
| Q4 | 17,585 (25.0) | 563,695 (25.0) | 87,320 (15.0) | 63,855 (24.0) |
| Missing | 3,665 (5.2) | 142,455 (6.4) | 61,565 (10.0) | 19,625 (7.4) |
| **Origin** |  |  |  |  |
| Danish | 62,015 (88.0) | 1,969,725 (88.0) | 520,870 (88.0) | 233,935 (88.0) |
| non-Danish | 8,745 (12.0) | 263,905 (12.0) | 68,830 (12.0) | 31,490 (12.0) |
| Missing | 75 (0.1) | 2,730 (0.1) | - | - |
| **Reproductive factors** |  |  |  |  |

|  | **VB-affected**<br>**pregnancy,**<br>**N=70,835** | **VB-unaffected**<br>**pregnancy,**<br>**N=2,236,360** | **Termination,**<br>**N=589,700** | **Miscarriage,**<br>**N=265,430** |
| --- | --- | --- | --- | --- |
| **Distinct women** | **N=66,881** | **N=1,198,206** | **N=424,834** | **N=225,103** |
| **Characteristic** | **n (%)<sup>a</sup>** | **n (%)<sup>a</sup>** | **n (%)<sup>a</sup>** | **n (%)<sup>a</sup>** |
| Previous vaginal bleeding diagnosis irrespective of pregnancy end | 8,145 (11.0) | 92,455 (4.1) | 33,625 (5.7) | 29,665 (11.0) |
| Previous vaginal bleeding within 20 gw of a pregnancy ending in delivery | 3,990 (5.6) | 42,270 (1.9) | 17,300 (2.9) | 8,655 (3.3) |
| History of ≥ 1 termination | 16,250 (23.0) | 350,845 (16.0) | 170,700 (29.0) | 53,520 (20.0) |
| History of ≥ 1 miscarriage | 18,315 (26.0) | 269,910 (12.0) | 57,425 (9.7) | 41,290 (16.0) |
| Hypertensive disorders of pregnancy | 2,490 (3.5) | 73,355 (3.3) | 18,955 (3.2) | 10,150 (3.8) |
| Placenta praevia | 125 (0.2) | 4,255 (0.2) | 1,350 (0.2) | 610 (0.2) |
| Abruptio placentae | 445 (0.6) | 9,000 (0.4) | 3,075 (0.5) | 1,580 (0.6) |
| Gestational diabetes | 300 (0.4) | 11,495 (0.5) | 2,705 (0.5) | 1,660 (0.6) |
| Abruptio placentae in a current pregnancy | 870 (1.2) | 11,985 (0.5) | - | - |
| Placenta praevia in a current pregnancy | 635 (0.9) | 9,105 (0.4) | - | - |
| Gestational diabetes in a current pregnancy | 925 (1.3) | 27,265 (1.2) | - | - |
| Gestational age at delivery, weeks (median, 25-75 percentile) | 39 (38-40) | 40 (39-41) | - | - |
| **Chronic conditions** |  |  |  |  |
| CVD, thrombosis, or diabetes<sup>b</sup> | 1,850 (2.6) | 55,975 (2.5) | 11,450 (1.9) | 7,085 (2.7) |
| Obesity diagnosis | 2,760 (3.9) | 109,170 (4.9) | 12,600 (2.1) | 6,975 (2.6) |
| PCOS | 460 (0.6) | 12,935 (0.6) | 1,245 (0.2) | 1,335 (0.5) |
| Thyroid disorders | 1,215 (1.7) | 36,105 (1.6) | 6,980 (1.2) | 3,950 (1.5) |
| Rheumatic disorders | 45 (<0.1) | 935 (<0.1) | 260 (<0.1) | 130 (<0.1) |
| COPD | 725 (1.0) | 18,820 (0.8) | 5,020 (0.9) | 2,250 (0.8) |
| Chronic kidney diseases | 170 (0.2) | 3,985 (0.2) | 1,155 (0.2) | 555 (0.2) |
| Chronic liver diseases | 115 (0.2) | 3,590 (0.2) | 1,120 (0.2) | 460 (0.2) |
| Cancer | 315 (0.4) | 9,675 (0.4) | 2,460 (0.4) | 1,165 (0.4) |
| Schizophrenia<sup>c</sup> | 405 (0.6) | 9,725 (0.4) | 6,300 (1.1) | 1,570 (0.6) |
| Autism spectrum disorders<sup>c</sup> | 30 (<0.1) | 855 (<0.1) | 550 (<0.1) | 105 (<0.1) |
| Neurotic disorders<sup>c</sup> | 4,315 (6.1) | 100,970 (4.5) | 46,540 (7.9) | 13,720 (5.2) |
| Eating disorders<sup>c</sup> | 520 (0.7) | 14,620 (0.7) | 6,005 (1.0) | 1,750 (0.7) |
| Personality disorder<sup>c</sup> | 1,580 (2.2) | 34,715 (1.6) | 21,050 (3.6) | 5,505 (2.1) |
| Intellectual disability<sup>c</sup> | 35 (<0.1) | 850 (<0.1) | 685 (0.1) | 140 (<0.1) |
| Behavioral disorder<sup>c</sup> | 50 (<0.1) | 1,360 (<0.1) | 1,020 (0.2) | 215 (<0.1) |
| Alcohol or drug abuse<sup>c</sup> | 540 (0.8) | 11,400 (0.5) | 11,575 (2.0) | 2,505 (0.9) |
| Any psychiatric condition<sup>c</sup> | 6,020 (8.5) | 153,160 (6.8) | 67,350 (11.0) | 20,195 (7.6) |

<sup>a</sup> All counts are clouded to the nearest 5 to prevent the identification of individuals and adhere to Statistics Denmark requirements. Percentages ≥10% are rounded to the nearest whole number and percentages <10% are rounded to the first decimal place.
<sup>b</sup> Diabetes mellitus types 1 and 2, hypertension, deep vein thrombosis, pulmonary embolism, haemorrhagic or ischemic stroke, ischaemic heart disease, atrial fibrillation or flutter, heart failure or use of clopidogrel or statins (according to data availability).
<sup>c</sup> Data on psychiatric hospital encounters and outpatient specialist clinic encounters were available from 1995.
COPD, chronic obstructive pulmonary disease; CVD, cardiovascular diseases; PCOS, polycystic ovary syndrome

We identified primary and secondary diagnoses as well as procedure codes for pregnancy termination and miscarriage between 1979 and 2017 at all hospital encounters from the DNPR. Women with multiple records of identical diagnostic or procedure codes for pregnancy termination or miscarriage on different dates could re-enter the respective population of terminated or miscarried pregnancies 180 days after the previous episode, whereby it was considered to represent a different pregnancy. When multiple dates characterised a hospital-based encounter for pregnancy termination or miscarriage, the earliest of them was the index date. This algorithm was introduced to avoid including hospital visits possibly related to a previous termination or miscarriage complications. We initiated the follow-up from the date of the woman’s hospital encounter (Table 1, available as Supplementary data) for termination or miscarriage. The observations with missing age or age <12 years, unknown gestational age for pregnancies ending in childbirth, two or more reproductive events (delivery, miscarriage, termination) recorded on the same date, or personal identifiers not found in the CRS were excluded from the final study population (<2% of all eligible observations; Figure 1). We also ascertained a population of women at the end of the first identifiable pregnancy for a separate set of analyses. In this data subset, each pregnancy observation corresponded to one unique woman whose index date was the end of her first pregnancy.

**Figure 1.**
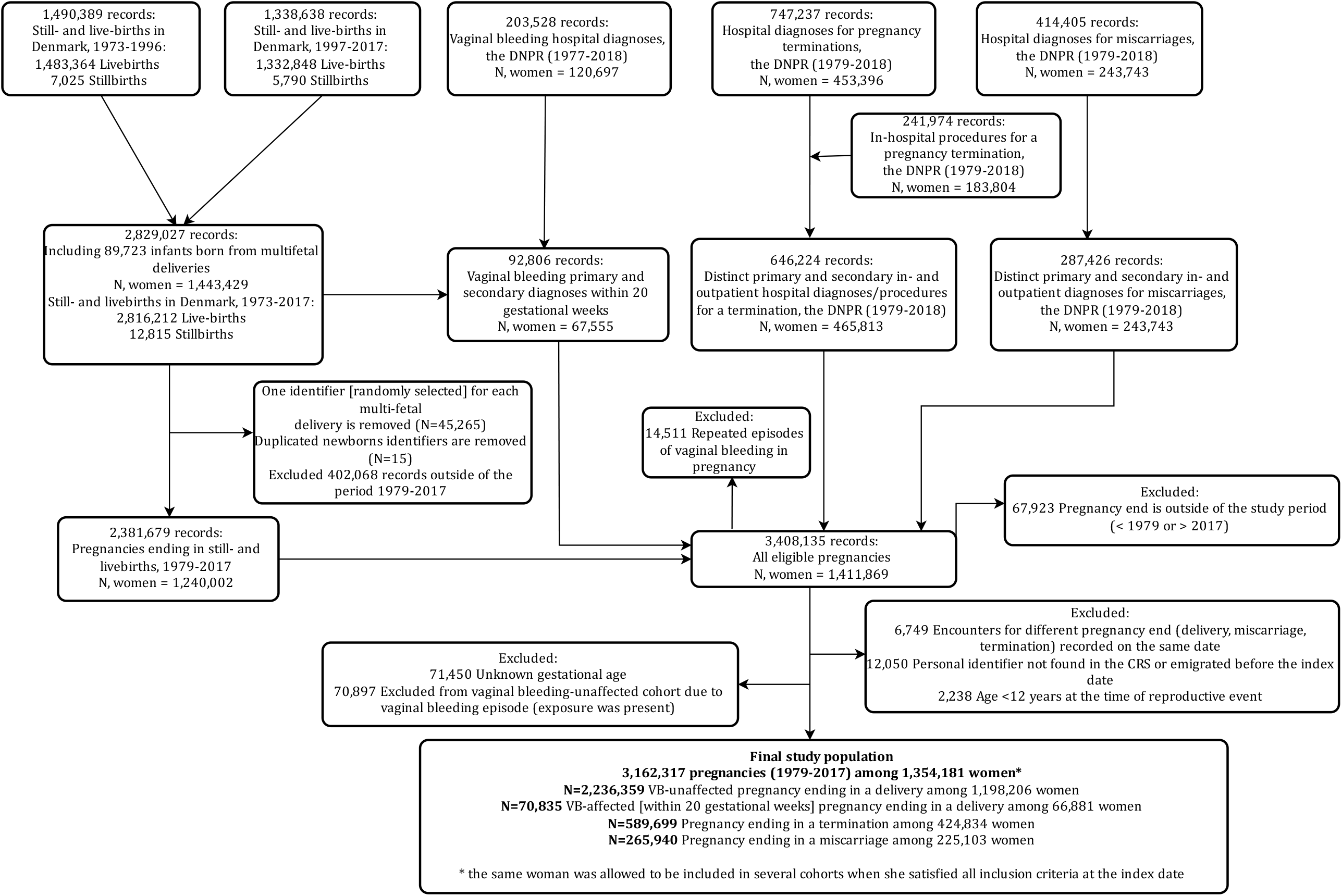
Flow chart depicting ascertainment of observations. Standalone figure

**Figure 2.**
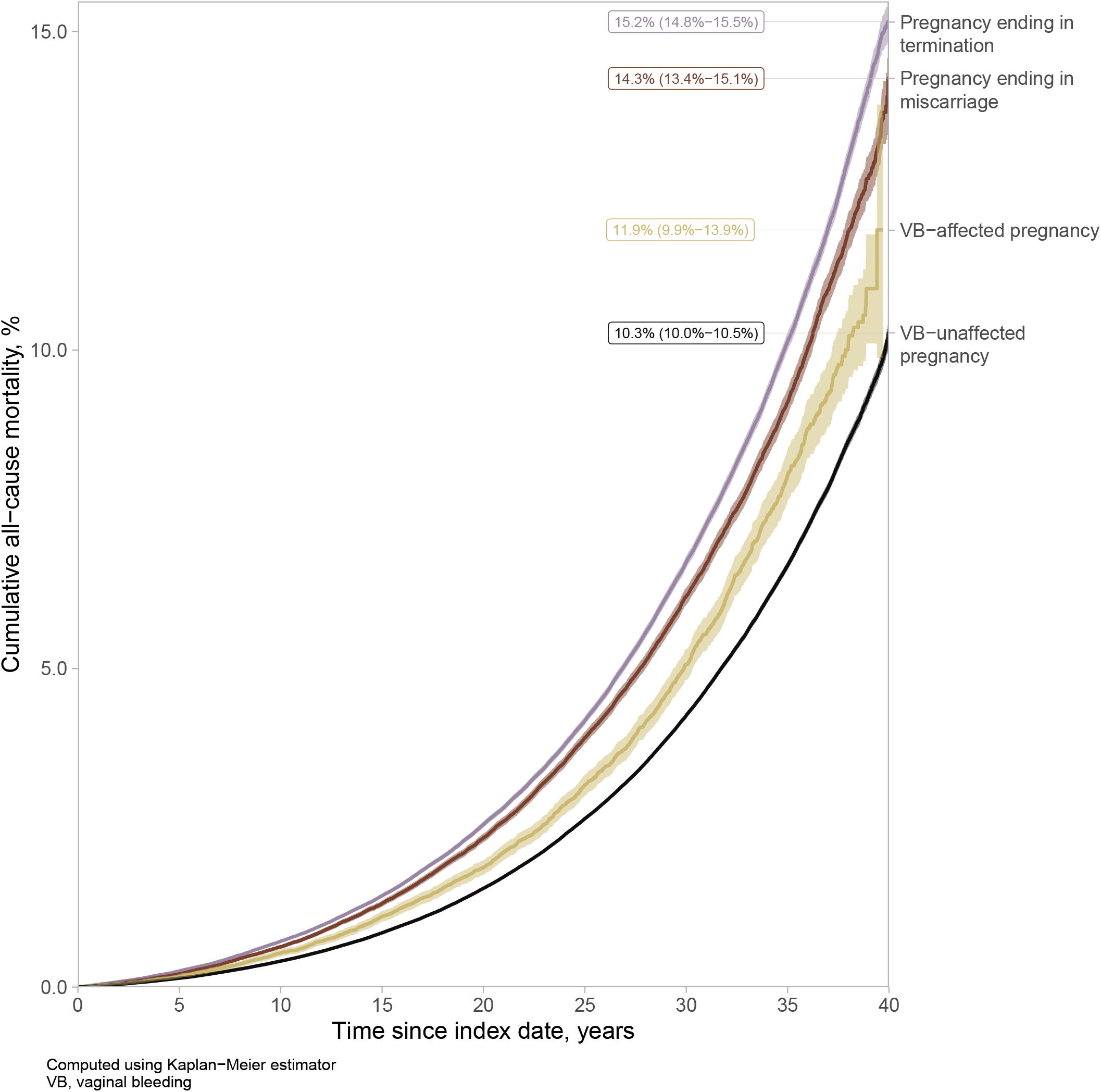
All-cause mortality following VB-affected and VB-unaffected pregnancies, pregnancies ending in a miscarriage, and pregnancies ending in a termination. Standalone figure

### Outcomes

We ascertained deaths from any cause using the vital status stored in the CRS.^21^ Specific causes of death were identified from the Danish Register of Causes of Death (DRCD),^32^ which records underlying, immediate, and several contributory causes of death along with the manner of death such as the death of natural causes, accident, suicide, or an act of violence. We used the underlying causes of death from the DRCD and classified them as natural causes when death was related to an underlying medical condition and as non-natural causes when the death occurred due to an accident, suicide, or violence. We further examined mortality due to several specific natural (cardiovascular disease, including stroke and myocardial infarction; any cancer; respiratory disease; endocrine, nutritional, and metabolic conditions, and nervous system disorders), non-natural (motor; non-motor vehicle accidents and violence; suicides), and all other causes (Table 1, available in the Supplementary data).

### Covariables

We used a directed acyclic graph to depict the relations between the variables in this study, both measured and unmeasured (Figure 1, available in the Supplementary data). We ascertained the following potential confounding factors and mortality risk factors before the index date: reproductive history (parity, number of previous pregnancies, history of at least one pregnancy termination or miscarriage, hypertensive disorders of pregnancy, placenta praevia, abruptio placentae), a woman’s age, calendar year at pregnancy end, socioeconomic factors (civil status, highest completed education level, employment status, and personal year-specific income in quartiles), history of chronic conditions (obesity, polycystic ovary syndrome, thyroid disorders, rheumatic disorders with heart involvement, chronic obstructive pulmonary disease, chronic liver and kidney diseases, hyperlipidemia, any cardiovascular or metabolic conditions [diabetes mellitus type 1 and 2, hypertension, deep vein thrombosis, pulmonary embolism, stroke, ischaemic heart disease, atrial fibrillation or flutter, heart failure], any cancer), and history of psychiatric conditions.

We ascertained women’s smoking status and BMI as potential confounders; these were only available for the subset of pregnancies ending in childbirth. For pregnancies with an index date after 1995, we ascertained the medication use history before the index date (non-steroid anti-inflammatory drugs including aspirin, steroids for systemic use, antiinfectives, antipsychotics, antidepressants, antiepileptics, and attention-deficit/hyperactivity disorder medication) as a proxy of healthcare-seeking behaviour and overall health beyond chronic conditions identified via hospital-based encounters (Table 1, available in the Supplementary data).

#### Statistical analyses

We followed the pregnancies until the earliest of death, emigration, or study end, on 31 December 2018 for all-cause mortality and 31 December 2017 for cause-specific mortality according to the last available data source update. We computed mortality rates per 10,000 person-years (PY) and cumulative mortality at 10, 20, 30, and 40 years of follow-up treating death from other causes and emigration as competing events (Table 2, available in the Supplementary data). We used the Kaplan-Meier estimator to construct cumulative incidence curves for all-cause mortality.^33,34^ We computed adjusted hazard ratios (HRs) with 95% confidence intervals (CIs) for all-cause and cause-specific mortality using cause-specific Cox proportional hazards regression with stabilized inverse probability of treatment weights (IPTWs).^35–37^ The weights were truncated at the 99^th^ percentile.^38,39^ The covariables included in the propensity score models were age at the pregnancy end, the calendar year of pregnancy end, reproductive history, socioeconomic factors, and pre-existing somatic and psychiatric conditions as described earlier. Analyses of pregnancies ending in childbirth were additionally adjusted for placenta-related conditions (hypertensive disorders of pregnancy including preeclampsia-eclampsia, placenta praevia, and abruptio placentae) in the current pregnancy. To allow comparability with earlier literature, we computed HRs adjusted for the same set of covariables as IPTW-adjusted models using multivariable Cox proportional hazards regression. Evaluation of log-minus-log plots showed no violations of hazards proportionality assumption for conducted analyses.

**Table 2.** Mortality rates following vaginal bleeding-affected and vaginal bleeding-unaffected pregnancies ending in childbirth and pregnancies ending in termination or miscarriage, 1979-2017/2018.

|  | No. of events |  |  |  | Mortality rate per 10,000 PY (95% CI) |  |  |  |
| --- | --- | --- | --- | --- | --- | --- | --- | --- |
|  | Exposure |  | Comparators |  | Exposure |  | Comparators |  |
|  | VB-affected pregnancy | VB-unaffected pregnancy | Termination | Miscarriage | VB-affected pregnancy | VB-unaffected pregnancy | Termination | Miscarriage |
| **All identifiable pregnancies<sup>a</sup>** | **N=70,835** | **N=2,236,360** | **N=589,700** | **N=265,430** |  |  |  |  |
| **Distinct women** | **N=66,881** | **N=1,198,206** | **N=424,834** | **N=225,103** |  |  |  |  |
| All-cause mortality<sup>b</sup> | 2,320 | 55,030 | 27,500 | 10,865 | 15.2 (14.6-15.9) | 12.7 (12.6-12.8) | 21.9 (21.6-22.1) | 19.2 (18.8-19.6) |
| Natural causes<sup>c</sup> | 1,545 | 37,250 | 17,830 | 7,160 | 10.6 (10.1-11.2) | 9.0 (8.9-9.1) | 14.8 (14.6-15.0) | 13.2 (12.9-13.5) |
| Cancer | 880 | 23,300 | 10,090 | 4,080 | 6.1 (5.7-6.5) | 5.6 (5.6-5.7) | 8.4 (8.2-8.6) | 7.5 (7.3-7.8) |
| Overall CVD | 210 | 4,835 | 2,425 | 940 | 1.4 (1.3-1.6) | 1.2 (1.1-1.2) | 2.0 (1.9-2.1) | 1.7 (1.6-1.9) |
| CVD other than myocardial infarction and stroke | 135 | 3,190 | 1,530 | 600 | 0.9 (0.8-1.1) | 0.8 (0.7-0.8) | 1.3 (1.2-1.3) | 1.1 (1.0-1.2) |
| Myocardial infarction | 35 | 780 | 410 | 140 | 0.2 (0.2-0.3) | 0.2 (0.2-0.2) | 0.3 (0.3-0.4) | 0.3 (0.2-0.3) |
| Stroke | 40 | 870 | 485 | 200 | 0.3 (0.2-0.4) | 0.2 (0.2-0.2) | 0.4 (0.4-0.4) | 0.4 (0.3-0.4) |
| Respiratory disease | 90 | 1,675 | 1,110 | 420 | 0.6 (0.5-0.8) | 0.4 (0.4-0.4) | 0.9 (0.9-1.0) | 0.8 (0.7-0.9) |
| Endocrine, nutritional, metabolic conditions | 35 | 705 | 390 | 190 | 0.2 (0.2-0.3) | 0.2 (0.2-0.2) | 0.3 (0.3-0.4) | 0.4 (0.3-0.4) |
| Nervous system condition | 50 | 1,180 | 655 | 225 | 0.4 (0.3-0.5) | 0.3 (0.3-0.3) | 0.5 (0.5-0.6) | 0.4 (0.4-0.5) |
| Non-natural causes<sup>c</sup> | 195 | 3,770 | 2,370 | 805 | 1.3 (1.2-1.5) | 0.9 (0.9-0.9) | 2.0 (1.9-2.1) | 1.5 (1.4-1.6) |
| Motor vehicle accident | 40 | 935 | 380 | 140 | 0.3 (0.2-0.4) | 0.2 (0.2-0.2) | 0.3 (0.3-0.3) | 0.3 (0.2-0.3) |
| Non-motor vehicle accident or violence | 85 | 1,450 | 1,125 | 380 | 0.6 (0.5-0.7) | 0.4 (0.3-0.4) | 0.9 (0.9-1.0) | 0.7 (0.6-0.8) |
| Suicide | 70 | 1,290 | 835 | 265 | 0.5 (0.4-0.6) | 0.3 (0.3-0.3) | 0.7 (0.6-0.7) | 0.5 (0.4-0.6) |
| All other causes | 275 | 5,640 | 3,195 | 1,325 | 1.9 (1.7-2.1) | 1.4 (1.3-1.4) | 2.7 (2.6-2.8) | 2.4 (2.3-2.6) |
| **First pregnancy of a woman** | **N=19,631** | **N=841,096** | **N=251,450** | **N=94,086** |  |  |  |  |
| **Distinct women** | **N=19,631** | **N=841,096** | **N=251,450** | **N=94,086** |  |  |  |  |
| All-cause mortality<sup>b</sup> | 530 | 17,935 | 13,385 | 4,440 | 12.9 (11.9-14.1) | 10.9 (10.8-11.1) | 22.5 (22.1-22.9) | 20.4 (19.8-21.0) |
| Natural causes<sup>c</sup> | 350 | 12,125 | 9,055 | 3,025 | 8.9 (8.1-9.9) | 7.8 (7.6-7.9) | 15.8 (15.5-16.2) | 14.4 (13.9-14.9) |
| Cancer | 200 | 7,555 | 5,165 | 1,730 | 5.1 (4.4-5.9) | 4.8 (4.7-5.0) | 9.0 (8.8-9.3) | 8.3 (7.9-8.7) |
| Overall CVD | 40 | 1,570 | 1,260 | 370 | 1.1 (0.8-1.4) | 1.0 (1.0-1.1) | 2.2 (2.1-2.3) | 1.8 (1.6-1.9) |
| CVD other than myocardial infarction and stroke | 30 | 1,045 | 785 | 220 | 0.7 (0.5-1.0) | 0.7 (0.6-0.7) | 1.4 (1.3-1.5) | 1.0 (0.9-1.2) |
| Myocardial infarction | 5 | 235 | 215 | 60 | 0.1 (0.0-0.3) | 0.1 (0.1-0.2) | 0.4 (0.3-0.4) | 0.3 (0.2-0.4) |
| Stroke | 10 | 285 | 260 | 90 | 0.2 (0.1-0.4) | 0.2 (0.2-0.2) | 0.4 (0.4-0.5) | 0.4 (0.3-0.5) |

Women's mortality following pregnancy affected by vaginal bleeding due to threatened miscarriage: a Danish cohort study
|  | No. of events |  |  |  | Mortality rate per 10,000 PY (95% CI) |  |  |  |
| --- | --- | --- | --- | --- | --- | --- | --- | --- |
|  | Exposure |  | Comparators |  | Exposure |  | Comparators |  |
|  | VB-affected pregnancy | VB-unaffected pregnancy | Termination | Miscarriage | VB-affected pregnancy | VB-unaffected pregnancy | Termination | Miscarriage |
| Respiratory disease | 20 | 490 | 610 | 200 | 0.5 (0.3-0.7) | 0.3 (0.3-0.3) | 1.1 (1.0-1.1) | 0.9 (0.8-1.1) |
| Endocrine, nutritional, metabolic conditions | 15 | 245 | 210 | 75 | 0.3 (0.2-0.6) | 0.2 (0.1-0.2) | 0.4 (0.3-0.4) | 0.3 (0.3-0.4) |
| Nervous system conditions | 10 | 395 | 325 | 115 | 0.3 (0.1-0.5) | 0.2 (0.2-0.3) | 0.6 (0.5-0.6) | 0.6 (0.5-0.7) |
| Non-natural causes<sup>c</sup> | 45 | 1,285 | 1,045 | 310 | 1.2 (0.9-1.6) | 0.8 (0.8-0.9) | 1.8 (1.7-1.9) | 1.5 (1.3-1.6) |
| Motor vehicle accident | 10 | 325 | 180 | 50 | 0.2 (0.1-0.4) | 0.2 (0.2-0.2) | 0.3 (0.3-0.4) | 0.2 (0.2-0.3) |
| Non-motor vehicle accident or violence | 20 | 470 | 440 | 130 | 0.5 (0.3-0.8) | 0.3 (0.3-0.3) | 0.8 (0.7-0.8) | 0.6 (0.5-0.7) |
| Suicide | 20 | 455 | 410 | 120 | 0.5 (0.3-0.7) | 0.3 (0.3-0.3) | 0.7 (0.6-0.8) | 0.6 (0.5-0.7) |
| All other causes | 65 | 1,905 | 1,505 | 545 | 1.7 (1.3-2.1) | 1.2 (1.2-1.3) | 2.6 (2.5-2.8) | 2.6 (2.4-2.8) |

<sup>a</sup> All counts are rounded to the nearest 5 to prevent the identification of individuals
<sup>b</sup> Data were available through 2018
<sup>c</sup> Data were available through 2017
CI, Confidence interval; CVD, Cardiovascular diseases; VB, Vaginal bleeding; PY, person-years

We conducted analyses of all identifiable pregnancies of a woman. In these analyses, we estimated robust standard errors to account for repeated observations of the same woman.^34,40^ We did not stop the follow-up of the observations at the subsequent pregnancy occurrence to avoid informative censoring. We accounted to the reproductive history and history of previous exposure at each subsequent pregnancy of a woman. In a separate set of analyses, we used only the first identifiable pregnancy of a woman.

We conducted several sensitivity analyses. First, we used a data subset of all pregnancies with the index date in 1994-2017 and the most complete health data while additionally controlling for the history of medication use before the index date as a proxy of overall women’s health. In the analyses of pregnancies ending in a childbirth, we additionally adjusted for smoking status. Second, we investigated all-cause, natural and non-natural causes of mortality among women whose last identifiable pregnancy was recorded at the age of 35 years or later.

We used RStudio^41^ and R^42^ 3.6.0-4.1.0 with the following packages: tidyverse,^43^ survival,^34^, cmprsk,^44^ epiR^45^, and WeightIt^39^ for statistical analyses. Data were accessed, managed, and analysed on Statistics Denmark servers.^22^ To prevent the identification of individuals, counts were masked to comply with Statistics Denmark data security rules.

This study was conducted according to Danish legislation, which does not require individuals’ consent or ethical approval for registry-based research. This study was reported to the Danish Data Protection Agency^46^ and registered at Aarhus University with project nr. 2016-051-000001, sequential number 605.

## Results

### Descriptive characteristics

Among 1,354,181 women, we identified 3,162,317 eligible pregnancies between 1979 and 2017. Of these, 2.2% (N=70,835) were pregnancies VB-affected within 20 gestational weeks and ending in childbirth, 70.7% (N=2,236,359) were pregnancies VB-unaffected within 20 gestational weeks and ending in childbirth, 18.7% (N=589,969) were pregnancies ending in a termination, and 8.4% (N=265,940) pregnancies ending in a miscarriage. Age distribution was similar for women with VB-affected and VB-unaffected pregnancies and pregnancies ending in a miscarriage, while pregnancies ending in a termination were more prevalent in younger women. Women with pregnancies ending in a termination were less likely to be married or employed, to have completed higher education, or to belong to the highest year-specific income quartile than women with pregnancies ending in a childbirth or a miscarriage (Table 1). Women with a VB-affected pregnancy were as likely as women with a miscarriage to have a history of VB (11.0%). Compared with VB-unaffected pregnancies (6.8%), VB-affected pregnancies (8.5%) and pregnancies ending in a termination (11.0%) were more likely to be of a woman with a history of any psychiatric condition. In analyses of all identifiable pregnancies, the history of at least one miscarriage was most prevalent among VB-affected pregnancies (26.0%) and in pregnancies ending in a miscarriage (16.0%). Chronic comorbidities were rare and similarly distributed across the cohorts (Table 1).

### All-cause and cause-specific mortality

The median follow-up for all pregnancies was 20 years (25th-75th percentile, 10.8-28.9 years). At the end of follow-up, there were 2,320 deaths from any cause following VB-affected pregnancy (mortality rate: 15.2, 95% CI: 14.6-15.9 per 10,000 PY), 55,030 deaths following VB-unaffected pregnancy (mortality rate: 12.7, 95% CI: 12.6-12.8), 27,500 deaths following a termination (mortality rate: 21.9, 95% CI: 21.6-22.1) and 10,865 deaths following a miscarriage (mortality rate: 19.2, 95% CI: 18.8-19.6). Mortality rates per 10,000 PY for death from natural causes were 10.6 (95% CI: 10.1-11.1) following VB-affected pregnancy and 9.0, 95% CI: 8.9-9.1 following VB-unaffected pregnancy. Mortality rate from natural causes was 14.8 (95% CI: 14.6-15.0) per 10,000 PY following a termination and 13.2 (95% CI: 12.9-13.5) per 10,000 PY following a miscarriage (Table 2). Results were similar in a subset restricted to women with the last identifiable delivery at the age of 35 years or later (Table 3, available in the Supplementary data). Irrespective of the type of pregnancy end, mortality from any cancer was the leading natural cause of death, followed by cardiovascular mortality and mortality due to respiratory conditions (Table 2). These patterns were consistent when we used a subset of the first identifiable pregnancy of a woman.

**Table 3.** Hazard ratios with 95% confidence intervals for the associations between vaginal bleeding and all-cause and cause-specific mortality, 1979-2017/2018.

|  |  | HRs (95% CI)<sup>a</sup> |  |  |
| --- | --- | --- | --- | --- |
|  |  | VB-affected vs VB-unaffected pregnancy | VB-affected pregnancy vs termination | VB-affected pregnancy vs miscarriage |
| **All-cause mortality<sup>b</sup>** | All identifiable pregnancies | 1.14 (1.09-1.19) | 0.85 (0.81-0.90) | 0.85 (0.80-0.90) |
|  | First pregnancy | 1.19 (1.09-1.30) | 0.83 (0.75-0.92) | 0.88 (0.80-0.97) |
| **Cause-specific mortality: natural causes<sup>c</sup>** | All identifiable pregnancies | 1.15 (1.09-1.22) | 0.86 (0.80-0.92) | 0.88 (0.82-0.95) |
|  | First pregnancy | 1.17 (1.05-1.31) | 0.80 (0.71-0.91) | 0.86 (0.77-0.97) |
| Cancer | All identifiable pregnancies | 1.07 (1.00-1.15) | 0.82 (0.75-0.89) | 0.90 (0.81-0.99) |
|  | First pregnancy | 1.09 (0.95-1.26) | 0.73 (0.61-0.87) | 0.85 (0.72-0.99) |
| Overall CVD | All identifiable pregnancies | 1.15 (0.99-1.34) | 0.84 (0.69-1.01) | 0.82 (0.67-1.01) |
|  | First pregnancy | 1.04 (0.76-1.43) | 0.70 (0.48-1.00) | 0.80 (0.56-1.13) |
| CVD other than myocardial infarction and stroke | All identifiable pregnancies | 1.11 (0.93-1.33) | 0.83 (0.67-1.04) | 0.83 (0.64-1.07) |
|  | First pregnancy | 1.06 (0.72-1.55) | 0.77 (0.50-1.19) | 0.90 (0.59-1.37) |
| Myocardial infarction | All identifiable pregnancies | 1.15 (0.80-1.65) | 0.97 (0.63-1.49) | 0.98 (0.59-1.65) |
|  | First pregnancy | 0.85 (0.35-2.11) | 0.74 (0.28-1.92) | 0.79 (0.31-2.04) |
| Stroke | All identifiable pregnancies | 1.29 (0.93-1.80) | 0.73 (0.49-1.08) | 0.70 (0.43-1.13) |
|  | First pregnancy | 1.15 (0.56-2.36) | 0.42 (0.17-1.04) | 0.54 (0.25-1.16) |
| Respiratory disease | All identifiable pregnancies | 1.53 (1.22-1.92) | 1.02 (0.78-1.34) | 0.99 (0.72-1.36) |
|  | First pregnancy | 1.68 (1.03-2.72) | 0.97 (0.59-1.60) | 0.90 (0.54-1.49) |
| Endocrine, nutritional, metabolic conditions | All identifiable pregnancies | 1.29 (0.89-1.86) | 1.16 (0.77-1.77) | 0.96 (0.61-1.52) |
|  | First pregnancy | 1.95 (1.11-3.44) | 1.45 (0.75-2.79) | 1.12 (0.60-2.12) |
| Nervous system condition | All identifiable pregnancies | 1.19 (0.89-1.61) | 0.75 (0.52-1.08) | 1.06 (0.71-1.60) |
|  | First pregnancy | 0.97 (0.52-1.79) | 0.66 (0.33-1.29) | 0.66 (0.35-1.24) |
| **Cause-specific mortality: non-natural causes<sup>c</sup>** | All identifiable pregnancies | 1.27 (1.08-1.48) | 0.86 (0.71-1.03) | 0.90 (0.73-1.11) |
|  | First pregnancy | 1.37 (1.02-1.85) | 1.09 (0.79-1.50) | 1.10 (0.79-1.52) |
| Motor vehicle accident | All identifiable pregnancies | 1.00 (0.71-1.41) | 1.04 (0.70-1.56) | 0.93 (0.57-1.50) |
|  | First pregnancy | 1.02 (0.51-2.03) | 1.32 (0.65-2.70) | 1.21 (0.58-2.53) |
| Non-motor vehicle accidents or violence | All identifiable pregnancies | 1.43 (1.13-1.81) | 0.83 (0.63-1.09) | 0.81 (0.58-1.12) |
|  | First pregnancy | 1.52 (0.96-2.43) | 1.11 (0.67-1.84) | 1.14 (0.69-1.87) |
| Suicide | All identifiable pregnancies | 1.36 (1.05-1.76) | 0.84 (0.62-1.14) | 1.10 (0.78-1.56) |
|  | First pregnancy | 1.58 (0.98-2.54) | 1.00 (0.59-1.69) | 1.08 (0.64-1.82) |
| All other causes | All identifiable pregnancies | 1.35 (1.18-1.53) | 0.91 (0.78-1.06) | 0.78 (0.65-0.94) |
|  | First pregnancy | 1.39 (1.08-1.80) | 1.00 (0.75-1.33) | 0.94 (0.71-1.25) |

<sup>a</sup> Analyses were adjusted using IPTW for age at the pregnancy end, calendar year of pregnancy, reproductive history, and pre-existing comorbidities (obesity, thyroid disorders, chronic obstructive pulmonary disease, chronic liver and kidney disease, hyperlipidaemia, cardiovascular, metabolic diseases), and psychiatric conditions. Analyses of pregnancies ending in a childbirth were additionally adjusted for placenta-related conditions at the index pregnancy.
<sup>b</sup> Data were available through 2018
<sup>c</sup> Data were available through 2017
CI, Confidence interval; CVD, cardiovascular diseases; IPTW, inverse probability of treatment weighting

All-cause mortality risks were similar for women with VB-affected and VB-unaffected pregnancy throughout the follow-up of up to 40 years and were marginally increased for the VB-affected cohort at the end of follow-up (11.9% vs 10.3%, respectively). Following a pregnancy ending in a termination or miscarriage, women’s all-cause mortality risk at the end of follow-up was 15.2% and 14.3%, respectively. At the end of follow-up, the cumulative mortality of natural causes was 8.2% for women with VB-affected pregnancy and 7.2% for women with VB-unaffected pregnancy; the cumulative mortality of non-natural causes was 0.5% following VB-affected and 0.3% following VB-unaffected pregnancy (Table 2, available in the Supplementary data).

### Hazard ratios for the associations between vaginal bleeding in pregnancy and women’s all-cause and cause-specific mortality

In the main IPTW-adjusted analyses, contrasts of VB-affected vs VB-unaffected pregnancies resulted in a HR (95% CI) for all-cause mortality of 1.14 (1.09-1.19). However, in the sensitivity analysis using the data subset with more data on potential confounders starting from 1994 onwards, the HR for all-cause mortality following VB-affected vs VB-unaffected pregnancy was 1.00, 95% CI: 0.90-1.12. Similarly, associations with mortality from natural causes (1.15, 95% CI: 1.09-1.22), including cardiovascular diseases overall (1.15, 95% CI: 0.99-1.34), stroke (1.29, 95% CI: 0.93-1.80), myocardial infarction (1.15, 95% CI: 0.80-1.65), and nervous system conditions (1.19, 95% CI: 0.89-1.61) attenuated in the sensitivity analyses (Table 4). Associations with mortality from respiratory disease and endocrine, nutritional, and metabolic conditions were not attenuated in the sensitivity analyses, but the estimates were imprecise. There was no evidence for an association of VB-affected pregnancy with mortality from cancer in either main (HR: 1.07, 95% CI: 1.00-1.15) or sensitivity analyses. An association between VB-affected pregnancy and mortality from non-natural causes (1.27, 95% CI: 1.08-1.48) also attenuated in the sensitivity analysis (1.04, 95% CI: 0.72-1.51). There was no association with death in motor vehicle accidents (1.00, 95% CI: 0.71-1.41), while associations with mortality from non-motor vehicle accidents or violence and with suicide meaningfully reduced in magnitude or disappeared in the sensitivity analysis (Table 4). Comparisons of VB-affected pregnancies ending in childbirth with pregnancies ending in a termination or miscarriage showed slightly decreased risks of all-cause and natural causes mortality, including cancer and cardiovascular mortality. The HRs for non-natural mortality, in general, were compatible with no association for comparisons of VB-affected pregnancy vs pregnancy ending in a termination or miscarriage (Table 4).

**Table 4.**
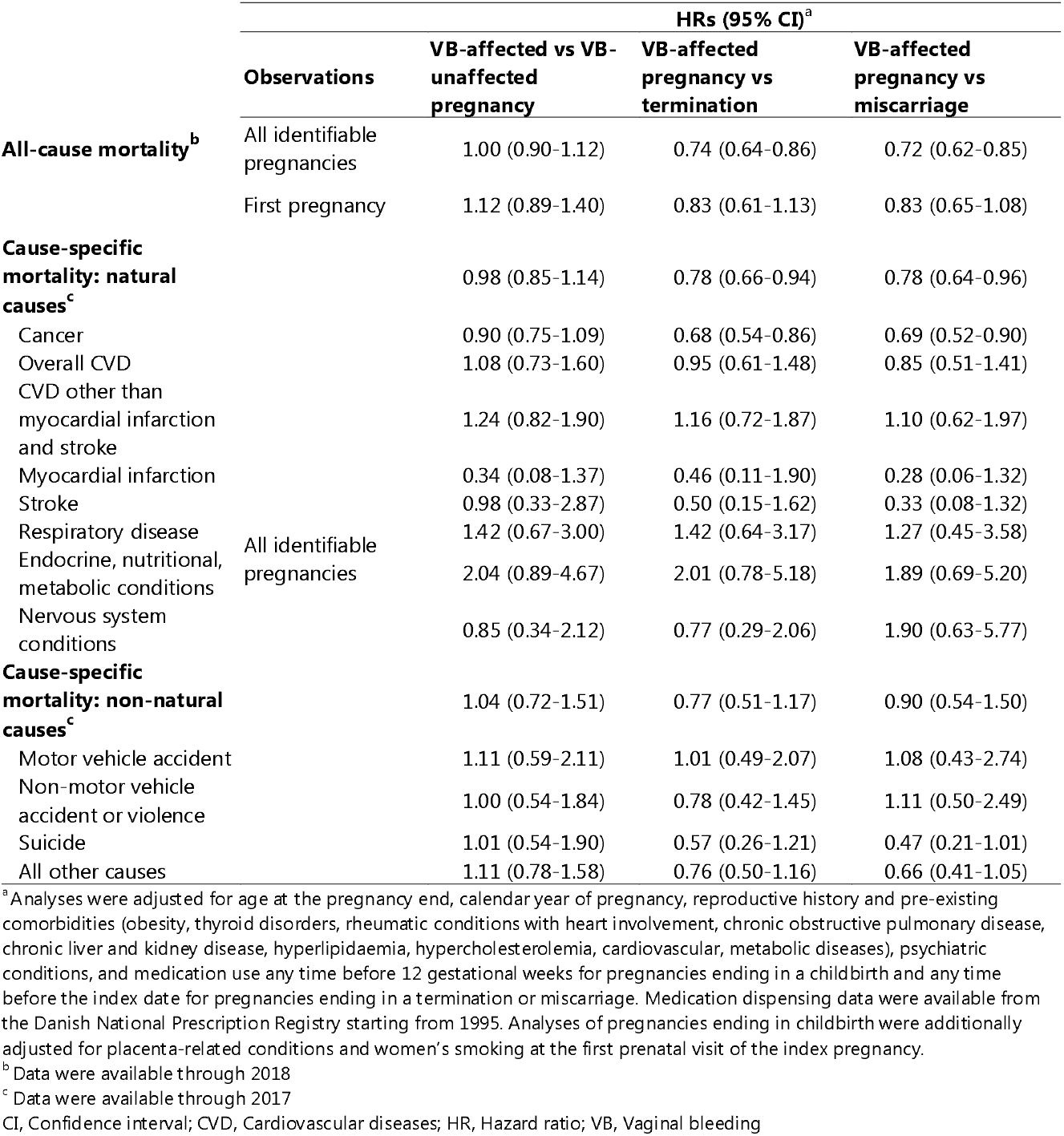
Hazard ratios with 95% confidence intervals for all-cause mortality and mortality from natural and non-natural causes among women with pregnancies ending in 1994-2017/2018.

The associations in the analyses of the first identifiable pregnancy of a woman were similar in magnitude to and less precise than the estimates from main analyses of all identifiable pregnancies (Table 3). Conventional multivariable Cox proportional hazards regression for adjusted analyses was in agreement with IPTW-adjusted results (Table 4, available in the Supplementary data). The analyses of the last identifiable pregnancy in women aged 35 years or later did not suggest a pattern of increased mortality risks associated with VB-affected pregnancy vs VB-unaffected pregnancy or remaining comparators (Table 5, available in the Supplementary data).

## Discussion

In this registry-based cohort study, we found no strong evidence of an increased risk of all-cause mortality, mortality from natural or non-natural causes following pregnancies affected by VB within 20 gestational weeks when compared with VB-unaffected pregnancies. The 15-27% increased risks of all-cause and cause-specific mortality attenuated towards the null value in the sensitivity analyses with more complete recent data and more strenuous confounding control. When contrasted with a termination or a miscarriage, VB-affected pregnancy ending in a childbirth was associated with a 15% reduced all-cause mortality and 10-17% reduced mortality risk from natural causes in main and sensitivity analyses; however, the residual confounding is of concern. HRs for non-natural causes of mortality were close to the null value with a miscarriage cohort as a comparator.

Previous studies of sequelae of VB in pregnancy mostly focused on short-term outcomes of newborns and mothers.^5^ This is the first study to specifically investigate the long-term mortality following VB-affected pregnancy ending in a childbirth. All-cause mortality rates in the present study were similar to previously reported mortality among women who gave birth.^47^

We conducted a registry-based study with essentially complete capture of deliveries in Denmark^23^ and women’s follow-up until death or emigration in routinely collected data at an individual level. The risk of selection bias due to differential loss-to-follow-up is expected to be low in this study. Of all pregnancies ending in childbirth, the prevalence of VB-affected pregnancies was 3% in this study. Other research suggests VB prevalence of up to 20-30% among all clinically recognised pregnancies.^48–53^ In a Danish study, 19% of women self-reported having had a VB during pregnancy, and 14% of these women had a hospital admission due to VB. This results in a point prevalence of 3-4% of hospital admissions due to VB among all pregnancies, which is consistent with our data.^48^ Although VB diagnosis due to threatened miscarriage was not validated in the DNPR, we expect it to be accurate since most pregnant women receive an ultrasound to establish foetal heartbeat upon VB presentation.^48^ We assume few false positives among VB-affected pregnancies are present. In support of that assumption, the records of miscarriage diagnoses in the DNPR have a positive predictive value (PPV) of >97%.^54^

The vital status from the CRS used for all-cause mortality ascertainment has very high accuracy and is continuously updated.^20,21,55^ Cause-specific mortality may be misclassified in a non-differential manner in regard to pregnancy status. Autopsies are not routinely performed in Denmark.^32,56^ One Danish study found 30% of the causes of death were different from the ones initially recorded after autopsies were conducted.^32,57^ If there was a true non-null association between VB-affected childbirths and subsequent increased women’s risk of cardiovascular death in comparison with VB-unaffected childbirths, non-differential misclassification could explain the null finding in the sensitivity analyses.

Lifestyle characteristics implicated as risk factors of both VB^58–60,6^ and mortality were smoking^61–63^ and BMI;^64,65^ however, their availability in this study was limited to pregnancies ending in childbirth. Although BMI was not a major confounding factor according to our data, attenuation of HRs in sensitivity analyses may partly be explained by more efficient confounding control in 1994-2017 calendar period and additional adjustment for comedications and smoking. In support of that, according to E-value analysis,^66^ a confounder or a set of confounders leading to bias in the same direction and associated 1.7 to 1.8-fold with the exposure and the outcome beyond the measured covariables would explain the associations with cause-specific mortality in analyses of VB-affected vs VB-unaffected pregnancies in 1979-2017. We did not model the joint effect of VB-affected pregnancy ending in a childbirth and miscarriages of subsequent pregnancies. We, thus, cannot rule out that the associations for the contrast of VB-affected vs VB-unaffected pregnancies found in main analyses in 1979-2017 can be explained by development of post-traumatic stress disorder, depression, anxiety following the occurrence of a miscarriage of a subsequent pregnancy.^4^ Other explanations for estimates’ attenuation to the null value in a sensitivity analysis is too few accrued events to detect associations. For comparisons of women with VB-affected pregnancy vs termination, the residual confounding by reproductive and socioeconomic factors beyond education, employment, and woman’s income at baseline is possible and may explain HRs below the null value for all-cause and cause-specific mortality.

## Conclusion

In this study, we found no evidence of increased all-cause or cause-specific mortality following vaginal bleeding-affected pregnancy when compared with pregnancies unaffected by vaginal bleeding or pregnancies interrupted by termination or miscarriage.

## Data Availability

Individual-level data used in this study are not publicly available in accordance with Danish legislation.

## Supplementary data

Supplementary data are available at *IJE* online.

## Contributors

ED, EHP, HTS, VE participated in designing the study. HTS organized and facilitated the data application. ED designed the statistical analysis plan under supervision of VE and EHP. ED analysed the data, EPH provided expertise and supervision in data analyses. All authors participated in the discussion of the findings and critical interpretation of the results. ED drafted the manuscript and led the writing. All authors participated in the critical revision of the manuscript and approved its final version. HTS is the guarantor.

## Ethics approval

Not required for registry-based research in Denmark. This study was reported to the Danish Data Protection Agency and registered at Aarhus University with project nr. 2016-051-000001, sequential number 605.

## Data sharing

Personal data are not available for public sharing to protect the identity of participants according to Danish legislation. The data sharing in aggregated form as presented in this publication is permitted. Source data are accessible to authorised researchers after application to Statistics Denmark. The corresponding author (ED) affirms that the manuscript is an honest, accurate, and transparent account of the study being report; that no important aspects of the study have been omitted; and that any discrepancies from the study as planned and registered have been explained.

## Supplementary data

Table 1 Definitions of the exposure, outcomes, and covariables (used ICD-8, ICD-10, and ATC codes)

Figure 1 Directed acyclic graph (DAG) depicting the relationship between measured and unmeasured variables in the study

Table 2 Cumulative all-cause and cause-specific mortality at 5, 10, 20, 30, and 40 years since the start of the follow-up

Table 3 Mortality rates and cumulative mortality following women with the last identifiable pregnancy at 35 years or later

Table 4 Adjusted hazard ratios (95% CIs) for the associations between vaginal bleeding and all-cause and cause-specific mortality using multivariable proportional hazards Cox regression, 1979-2018

Table 5 Adjusted hazard ratios (95% CIs) for the associations between vaginal bleeding and all-cause and cause-specific mortality following the last identifiable pregnancy at 35 years or later, Denmark, 1979-2018

