## Supplementary data for "Women’s mortality following pregnancy affected by vaginal bleeding due to threatened miscarriage: a Danish cohort study"

### **Table 1 Definitions of the exposure, outcomes, and covariables (used ICD-8, ICD-10, and ATC codes)**

|  | **ICD-8 codes** | **ICD-10 codes** | **Procedure codes** | **ATC codes** |
| --- | --- | --- | --- | --- |
| **Exposure and comparators** | | | | |
| **Vaginal bleeding in pregnancy (threatened abortion)** | 632.39 | DO200 | - | - |
| **Miscarriage (spontaneous abortion)** | 643xx; 634xx;  645xx | O03; O021A | - | - |
| **Pregnancy termination (medical or surgical abortion)** | 640xx; 641xx; 642xx | O04 | 94520 (abortus provocatus medicamentalis; KLCH (termination of pregnancy) | - |
| **Covariables** | | | | |
| **Comorbidities** | | | | |
| **Obesity** | 277.99 | E66 | - | - |
| **PCOS** | 256.90 | E28.2 | - | - |
| **Thyroid disorders** | 240xx-246xx | E00-E07 | - | - |
| **Rheumatic diseases with heart involvement** | 390.99, 391.99, 392.99, 392.09, 393.00, 393.01 | I00.0-I02.9; I05.0-I09.9 | - | - |
| **COPD** | 490xx-493xx; 515xx-518xx | J40-J47; J60-J67;  J68.4; J70.1; J70.3;  J84.1; J92.0; J96.1;  J98.2; J98.3 | - | - |
| **Chronic kidney diseases** | 249.02, 250.02, 753.10-753.19, 582xx, 583xx, 584xx, 590.09, 593.20, 792xx | 249.02, 250.02, 753.10-753.19, 582, 583, 584, 590.09, 593.20, 792;  E10.2, E11.2, E14.2,  N03, N05, N11.0,  N14; N16, N18-N19,  N26.9, Q61.1-Q61.4 | - | - |
| **Liver disease** | 571xx | K70.0, K70.3, K71.7,  K73, K74, K76.0,  B18, I85 | - | - |
| **Cancer** | 140-209 | C00-C96 | - | - |
| **Hyperlipidemia** | 272.01, 272.08, 272.09 | E78.1-E78.5 | - | - |
| **Hypercholesterolemia** | 272.00 | E78.0 | - | - |
| **Preeclampsia superimposed on chronic hypertension** | N/A | O11 | - | - |
| **Gestational proteinuria** | 637.01, 637.02 | O12 | - | - |
| **Gestational hypertension** | 637.00 | O13 | - | - |
| **Preeclampsia** | 637.03, 637.04, 637.09 | O14 | - | - |
| **Eclampsia** | 637.19 | O15 | - | - |
| **Any hypertensive disorder of pregnancy** | 637.01, 637.02, 637.00, 637.03, 637.04, 637.09, 637.19 | O11- O15 | - | - |
| **Placenta praevia** | 651.0x; 651.1x; 651.2x; 651.3x | O44 | - | - |
| **Abruptio placentae** | 6321x; 6514x | O45 | - | - |
| **Gestational diabetes** | 634.74, 644.9x, Y6449 | O24.4, O24.9 | - | - |
| **Cardiovascular disease or diabetes history:** | | | | |
| **Diabetes mellitus type 1** (same codes are used for diabetes mellitus type 1 and 2 until 1987) | 249 | E10, O24.0 | - | A10A*  *The first redeemed dispensing defined the date of diabetes mellitus type 1 outcome if no prior diagnostic code for diabetes mellitus type 1 was recorded. Otherwise, the date associated with the diagnostic record for diabetes mellitus type 1 defined the date of the condition. |
| **Diabetes mellitus type 2** | 250 | E11, E891, G590, G632, G730A, G990C, H280, H360, I792A, M142, N083 | - | A10B  *The first redeemed dispensing defined the date of diabetes mellitus type 2 outcome if no prior diagnostic code for diabetes mellitus type 2 was recorded. Otherwise, the date associated with the diagnostic record for diabetes mellitus type 2 defined the date of the condition. |
| **Hypertension** | 400.09-404.99 | I10.0-I15.9 | - | C02A, C02B, C02C (alpha-blockers); C02DA, C02L, C03A, C03B, C03D, C03E, C03X, C07C, C07D, C08G, C09BA, C09DA, C09XA52 (non-loop diuretics); C02BD, C02DD, C02DG, C04, C05 (vasodilators); C07 (beta-blockers); C08, C07F, C09BB, C09DB (calcium channel blockers); C09 (RAS-acting agents: ACE inhibitors and ARBs)*  *The second of 2+ redeemed dispensings of different antihypertensive drug classes within 180 days of each other defined the date of hypertension outcome if no prior diagnostic code for hypertension was recorded. Otherwise, the date associated with the diagnostic record for hypertension defined the date of the covariable. |
| **Ischemic heart disease** | 410xx-414xx | I20-I25 | - | - |
| **Heart failure** | 427.09-427.19 | I110, I130, I132, I50 | - | - |
| **Ischemic stroke** | 433-434 | I63-I64 | - | - |
| **Haemorrhagic stroke** | 430-432 | I60, I61 | - | - |
| **Deep vein thrombosis** | 451.00 | I80.1-I80.3 | - | - |
| **Pulmonary embolism** | 450.99 | I26 | - | - |
| **Clopidogrel** | - | - | - | B01AC04 |
| **Statins** | - | - | - | C10AA, C10B |
| **Any psychiatric disorder** | 290-315 | F00-F99 | - | - |
| **Mood disorders** | 296.x9 (excluding 296.89), 298.09, 298.19, 300.49, 301.19 | F30-F39, F92.0 | - | - |
| **Schizophrenia** | 295.x9, 96.89, 297.x9, 98.29-298.99, 299.04, 299.05, 99.09, 301.83 | F20-29 | - | - |
| **Autism spectrum disorder** | 299.01-299.03 | F84.0-84.1, F84.5, F84.8-84.9 | - | - |
| **Neurotic disorders** | 300.x9 (Excluding 300.49), 305.x9, 305.68, 307.99 | F40-F48 | - | - |
| **Eating disorders** | 305.60, 306.50, 306.58, 306.59 | F50 | - | - |
| **Personality disorders** | 301.x9 (Excluding 301.19), 301.80, 301.81, 301.82, 301.84 | F60 | - | - |
| **Intellectual disabilities** | 311.xx, 312.xx, 313.xx, 314.xx, 315.xx | F70-F79 | - | - |
| **Behavioural disorders** | 306.x9, 308.0x | F90-F98 | - | - |
| **Alcohol or drug abuse** | 291.xx, 303.x9, 303.20, 303.28, 303.90, 294.39, 304x9; 577.10, 571.09, 571.10 | F10, F11-F19; Z72.1, T51.0, K86.0, G31.2, G62.1, G72.1, I42.6, K29.2, R78.0, Z71.4 | - | - |
| **Comedications (for sensitivity analysis in 1994-2018)** | | | | |
| **Antipsychotics** | - | - | - | N05A |
| **Mood disorders medication** | - | - | - | N06 |
| **NSAIDs** | - | - | - | M01A, N02BA |
| **Aspirin** | - | - | - | B01AC06 |
| **Thyroid drugs** | - | - | - | H03 |
| **Steroids for systemic use** | - | - | - | H02A, H02B |
| **Anti-infectives for systemic use** | - | - | - | J01, J02, J04, J09, J12 |
| **Antiepileptics** | - | - | - | N03A |
| **ADHD medication** | - | - | - | N06BA01, N06BA02, N06BA04, N06BA09, N06BA12 |
| **Outcome: cause-specific mortality** | | | | |
| **Underlying causes of death** | | | | |
| **Natural causes (related to a medical condition)** | 0000-7969 | A00-R99 | - | - |
| **Cancer** | 140-239 | C00-D48 | - | - |
| **Cardiovascular diseases (CVD)** | 390-459 | I00-I99 | - | - |
| **Myocardial infarction** | 410 | I21-I23 | - | - |
| **Stroke** | 431-434 | I61, I63-6I4 | - | - |
| **Respiratory conditions** | 460-519 | J00-J99 | - | - |
| **Endocrine/nutritional/metabolic conditions** | 240-279 | E00-E90 | - | - |
| **Diseases of the nervous system conditions** | 320-359 | F01-F03, G00-G99 | - | - |
| **Non-natural causes** | 8000-9999 | V01-Y98 | - | - |
| **Motor vehicle accidents** | 8100-8230 | V01-V89 | - | - |
| **Suicides** | 9500-9599 | X60-X84 | - | - |
| **Non-vehicle accidents/violence** | 8000-8079, 8250-9499, 9600-9999 | V90-V99, W00-X59, X59-Y89 | - | - |
| **Other natural and non-natural causes** | 0000-7969 excl. codes above & 8000-9999 excl. codes classified above | A00-R99 excl. codes above & V01-Y98 excl. codes classified above | - | - |

### **Figure 1 Directed acyclic graph (DAG) depicting the relationship between measured and unmeasured variables in the study**

**Abbreviations**: PREG, pregnancy; PREG_BMI_t0_, body-mass index at the index date/pregnancy (at time zero); PRE**-**t0**,** marks covariables measured before the index date (before time zero); TAB, threatened abortion; U_1_, an unmeasured shared cause for both miscarriage and pregnancy termination and the DEATH_t0+40yrs_; U_2_, an unlikely unmeasured factor increasing the risk of migration and DEATH_t0+40yrs_; FERTILITY is unmeasured (latent node)

**Exposure**: Vaginal-bleeding affected pregnancy within the first 20 gestational weeks ending in childbirth (PREG_childbirthTAB_t0_)

**Outcome**: All-cause and cause-specific mortality

Directed solid line arrows: causal relationship (including temporality, e.g. A → B means that A causes and precedes B)

Bi-directed dashed line arrows: associations confounded by the unmeasured common shared causes (A ⟷ B means A ←U→ B, where U is unobserved)

DAG tool: https://causalfusion.net/app^1^

Potential mediators are not depicted on the DAG since we did not perform dynamic modelling with time-varying exposure and time-varying confounding and did not examine the specific mediation pathways following the vaginal bleeding-affected pregnancy.

### **Average treatment effect computation using inverse probability of treatment weighting (IPTW):**

For confounding adjustment, we computed stabilized IPTW as the ratio of the probability of the observed exposure level in the numerator: $Pr[A=1]$for VB-affected childbirths and $1-Pr[A=1]$for comparator pregnancies (where A=1 denotes having vaginal bleeding within 20 weeks of pregnancy) and the conditional probability of having the actual exposure level given confounders and mortality risk factors ($Pr[A=1|L]$for VB-affected pregnancies ending in childbirth and $1-Pr[A=1|L]$ otherwise, where L denotes measured confounders and mortality predictors) in the denominator.^2–5^

Stabilized IPT weights for exposed: $Pr[A=1]/ Pr[A=1|L]$

Stabilized IPT weights for unexposed/comparisons: $1-Pr[A=1]/ 1-Pr[A=1|L]$

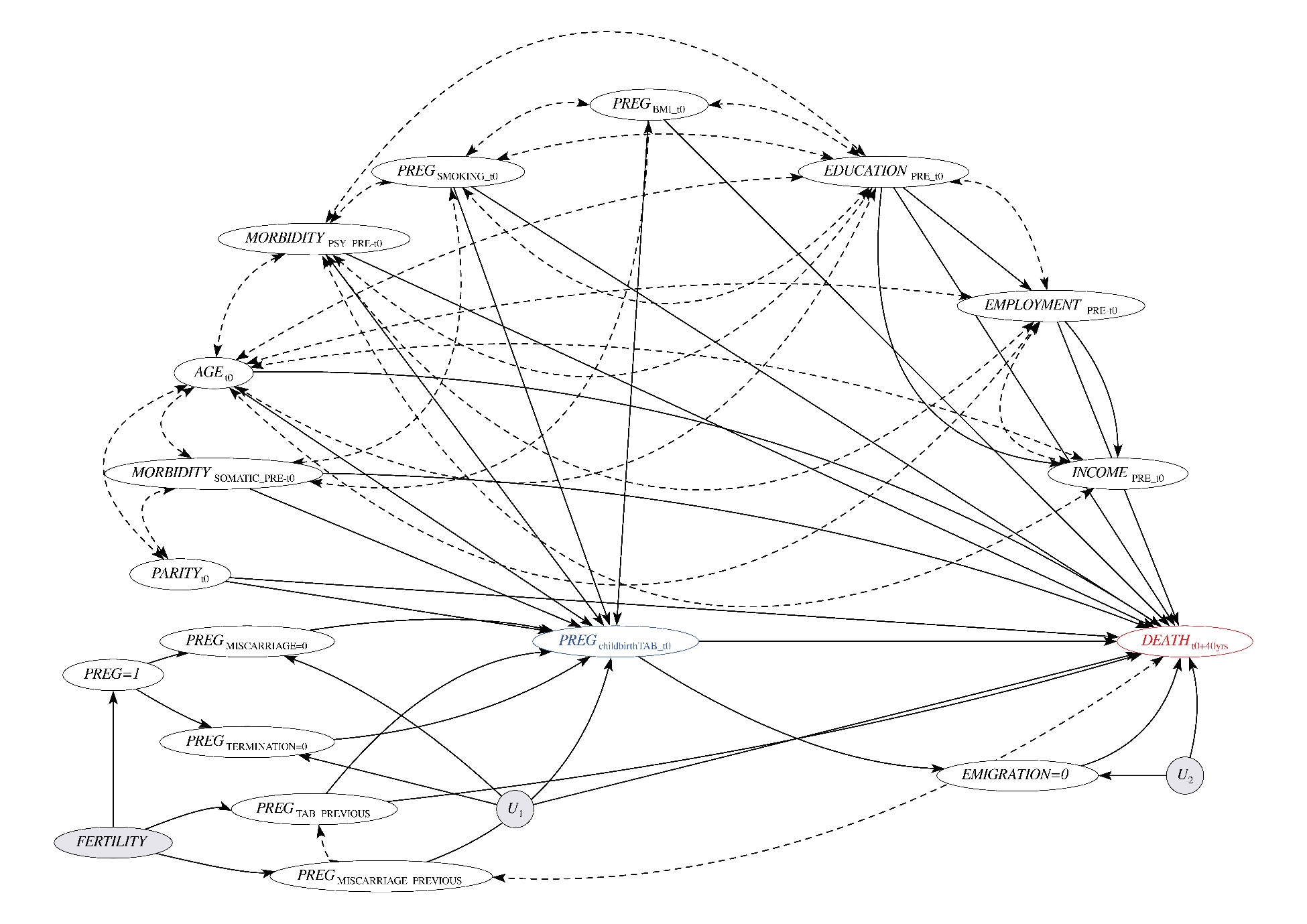

### **Table 2 Cumulative all-cause and cause-specific mortality at 5, 10, 20, 30, and 40 years since the start of the follow-up**

|  |  |  | **Cumulative risk (%)** | | | |
| --- | --- | --- | --- | --- | --- | --- |
|  |  |  | **Exposure** | **Comparators** | | |
|  | **Time, years** |  | **Vaginal bleeding-affected**  **pregnancy** | **Vaginal bleeding-unaffected**  **pregnancy** | **Termination** | **Miscarriage** |
| All-cause mortality^a^ | 5 | All identifiable pregnancies | 0.2 (0.1-0.2) | 0.1 (0.1-0.1) | 0.3 (0.2-0.3) | 0.2 (0.2-0.2) |
|  | 10 |  | 0.5 (0.5-0.6) | 0.4 (0.4-0.4) | 0.7 (0.7-0.7) | 0.6 (0.6-0.7) |
|  | 20 |  | 1.9 (1.8-2.0) | 1.6 (1.5-1.6) | 2.6 (2.5-2.6) | 2.3 (2.3-2.4) |
|  | 30 |  | 5.1 (4.8-5.3) | 4.3 (4.2-4.3) | 6.7 (6.6-6.8) | 6.1 (6.0-6.3) |
|  | 40 |  | 11.9 (9.9-13.9) | 10.3 (10.0-10.5) | 15.2 (14.8-15.5) | 14.3 (13.4-15.1) |
|  | 5 | First pregnancy | 0.1 (0.1-0.2) | 0.1 (0.1-0.1) | 0.2 (0.2-0.2) | 0.2 (0.2-0.2) |
|  | 10 |  | 0.4 (0.3-0.5) | 0.3 (0.3-0.3) | 0.6 (0.5-0.6) | 0.6 (0.5-0.6) |
|  | 20 |  | 1.4 (1.2-1.6) | 1.2 (1.2-1.3) | 2.2 (2.1-2.2) | 2.1 (2.0-2.2) |
|  | 30 |  | 4.5 (4.0-5.0) | 3.5 (3.4-3.6) | 5.9 (5.8-6.1) | 5.6 (5.4-5.8) |
|  | 40 |  | 9.9 (8.5-11.4) | 8.8 (8.6-9.1) | 14.2 (13.9-14.6) | 13.9 (12.9-14.9) |
| Natural causes^b^ | 5 | All identifiable pregnancies | 0.1 (0.1-0.1) | 0.1 (0.1-0.1) | 0.1 (0.1-0.1) | 0.1 (0.1-0.1) |
|  | 10 |  | 0.4 (0.3-0.4) | 0.3 (0.3-0.3) | 0.4 (0.4-0.4) | 0.4 (0.4-0.4) |
|  | 20 |  | 1.3 (1.2-1.4) | 1.1 (1.1-1.2) | 1.7 (1.7-1.8) | 1.7 (1.6-1.7) |
|  | 30 |  | 3.6 (3.4-3.9) | 3.2 (3.1-3.2) | 4.7 (4.6-4.8) | 4.4 (4.3-4.5) |
|  | 39 |  | 8.2 (7.4-9.1) | 7.2 (7.1-7.4) | 10.9 (10.6-11.2) | 9.9 (9.4-10.4) |
|  | 5 | First pregnancy | 0.1 (0.0-0.1) | 0.1 (0.1-0.1) | 0.1 (0.1-0.1) | 0.1 (0.1-0.1) |
|  | 10 |  | 0.3 (0.2-0.4) | 0.2 (0.2-0.2) | 0.3 (0.3-0.4) | 0.4 (0.3-0.4) |
|  | 20 |  | 1.0 (0.9-1.2) | 0.9 (0.9-0.9) | 1.6 (1.5-1.6) | 1.6 (1.5-1.7) |
|  | 30 |  | 3.1 (2.7-3.5) | 2.6 (2.5-2.6) | 4.4 (4.3-4.5) | 4.1 (4.0-4.3) |
|  | 39 |  | 7.4 (6.2-8.6) | 6.3 (6.1-6.5) | 10.3 (10.0-10.6) | 9.5 (8.9-10.0) |
| Non-natural causes^b^ | 5 | All identifiable pregnancies | 0.1 (0.0-0.1) | 0.0 (0.0-0.0) | 0.1 (0.1-0.1) | 0.1 (0.1-0.1) |
|  | 10 |  | 0.1 (0.1-0.2) | 0.1 (0.1-0.1) | 0.2 (0.2-0.2) | 0.2 (0.1-0.2) |
|  | 20 |  | 0.3 (0.2-0.3) | 0.2 (0.2-0.2) | 0.4 (0.4-0.4) | 0.3 (0.3-0.3) |
|  | 30 |  | 0.4 (0.3-0.4) | 0.3 (0.3-0.3) | 0.6 (0.5-0.6) | 0.4 (0.4-0.5) |
|  | 39 |  | 0.5 (0.4-0.6) | 0.3 (0.3-0.4) | 0.7 (0.6-0.8) | 0.5 (0.5-0.6) |
|  | 5 | First pregnancy | 0.0 (0.0-0.1) | 0.0 (0.0-0.0) | 0.1 (0.1-0.1) | 0.1 (0.0-0.1) |
|  | 10 |  | 0.1 (0.1-0.2) | 0.1 (0.1-0.1) | 0.2 (0.2-0.2) | 0.2 (0.1-0.2) |
|  | 20 |  | 0.3 (0.2-0.3) | 0.2 (0.2-0.2) | 0.4 (0.4-0.4) | 0.3 (0.3-0.4) |
|  | 30 |  | 0.4 (0.3-0.5) | 0.2 (0.2-0.3) | 0.5 (0.5-0.6) | 0.4 (0.4-0.5) |
|  | 39 |  | 0.4 (0.3-0.5) | 0.3 (0.3-0.3) | 0.7 (0.6-0.7) | 0.5 (0.5-0.6) |
| Overall CVD causes^b^ | 5 | All identifiable pregnancies | 0.0 (0.0-0.0) | 0.0 (0.0-0.0) | 0.0 (0.0-0.0) | 0.0 (0.0-0.0) |
|  | 10 |  | 0.0 (0.0-0.1) | 0.0 (0.0-0.0) | 0.1 (0.1-0.1) | 0.1 (0.1-0.1) |
|  | 20 |  | 0.2 (0.1-0.2) | 0.2 (0.2-0.2) | 0.3 (0.2-0.3) | 0.2 (0.2-0.3) |
|  | 30 |  | 0.5 (0.4-0.6) | 0.4 (0.4-0.4) | 0.6 (0.6-0.7) | 0.6 (0.5-0.6) |
|  | 39 |  | 0.9 (0.7-1.1) | 0.9 (0.9-1.0) | 1.5 (1.4-1.6) | 1.3 (1.1-1.5) |
|  | 5 | First pregnancy | 0.0 (0.0-0.0) | 0.0 (0.0-0.0) | 0.0 (0.0-0.0) | 0.0 (0.0-0.0) |
|  | 10 |  | 0.0 (0.0-0.0) | 0.0 (0.0-0.0) | 0.1 (0.0-0.1) | 0.1 (0.0-0.1) |
|  | 20 |  | 0.2 (0.1-0.3) | 0.1 (0.1-0.1) | 0.2 (0.2-0.2) | 0.2 (0.2-0.2) |
|  | 30 |  | 0.4 (0.2-0.5) | 0.3 (0.3-0.4) | 0.6 (0.6-0.7) | 0.5 (0.5-0.6) |
|  | 39 |  | 0.8 (0.4-1.1) | 0.8 (0.8-0.9) | 1.5 (1.4-1.6) | 1.2 (1.0-1.4) |
| ^a^ Data were available through 2018; cumulative risk computed using Kaplan-Meier estimator | | | | | | |
| ^b^ Data were available through 2017; cumulative risks computed using cumulative incidence function estimator | | | | | | |

### **Table 3 Mortality rates and cumulative mortality following women with the last identifiable pregnancy at 35 years or later**

|  | Exposure | Comparators | | |
| --- | --- | --- | --- | --- |
|  | VB-affected pregnancy^a^ | VB-unaffected pregnancy^a^ | Termination^a^ | Miscarriage^a^ |
|  | **No. of women at risk** | | | |
|  | 7,905 | 245,015 | 100,080 | 28,725 |
|  | **No. of events** | | | |
| All-cause mortality^b^ | 355 | 8,020 | 9,095 | 2,280 |
| Mortality from natural causes^c^ | 250 | 5,755 | 6,600 | 1,585 |
| Mortality from non-natural causes^c^ | 15 | 320 | 375 | 120 |
|  | **Mortality rate per 10,000 PY (95% CI)** | | | |
| All-cause mortality^b^ | 24.9 (22.4-27.6) | 22.0 (21.6-22.5) | 43.2 (42.3-44.1) | 43.1 (41.4-44.9) |
| Mortality from natural causes^c^ | 18.5 (16.4-20.9) | 16.9 (16.5-17.3) | 32.7 (31.9-33.5) | 31.5 (30.0-33.1) |
| Mortality from non-natural causes^c^ | 1.0 (0.6-1.7) | 0.9 (0.8-1.0) | 1.9 (1.7-2.1) | 2.3 (1.9-2.8) |
|  | **Cumulative all-cause mortality**^b^ | | | |
| **Time, years** |  |  |  |  |
| 5 | 0.2 (0.1-0.3) | 0.3 (0.3-0.3) | 0.4 (0.4-0.5) | 0.5 (0.4-0.6) |
| 10 | 0.9 (0.7-1.1) | 0.8 (0.8-0.9) | 1.3 (1.2-1.4) | 1.6 (1.4-1.7) |
| 20 | 3.5 (2.9-4.0) | 3.3 (3.2-3.4) | 4.9 (4.8-5.1) | 5.7 (5.3-6.0) |
| 30 | 10.2 (8.9-11.5) | 9.1 (8.9-9.3) | 12.9 (12.6-13.2) | 14.3 (13.6-15.0) |
| 40 | 20.4 (16.7-24.0) | 22.4 (20.6-24.1) | 29.4 (28.3-30.5) | 30.2 (27.9-32.3) |
| ^a^ Last recorded pregnancy in the Medical Birth Registry or the National Patient Registry by Dec 31, 2017  ^b^ Data were available through 2018  ^c^ Data were available through 2017 | | | | |

### **Table 4 Adjusted hazard ratios (95% CIs) for the associations between vaginal bleeding and all-cause and cause-specific mortality using multivariable (conventional) proportional hazards Cox regression, 1979-2018**

|  |  | **HRs (95% CI)**^a^ | | |
| --- | --- | --- | --- | --- |
|  |  | **VB-affected vs VB-unaffected pregnancy** | **VB-affected pregnancy vs termination** | **VB-affected pregnancy vs miscarriage** |
| **All-cause mortality^b^** | All identifiable pregnancies | 1.08 (1.04-1.13) | 0.85 (0.82-0.89) | 0.85 (0.80-0.90) |
|  | First pregnancy | 1.11 (1.02-1.21) | 0.92 (0.84-1.00) | 1.01 (0.92-1.10) |
| **Cause-specific mortality: natural causes^c^** | All identifiable pregnancies | 1.09 (1.04-1.15) | 0.92 (0.87-0.97) | 0.91 (0.85-0.97) |
|  | First pregnancy | 1.08 (0.97-1.20) | 0.94 (0.84-1.05) | 1.00 (0.90-1.12) |
| Cancer | All identifiable pregnancies | 1.03 (0.96-1.10) | 0.88 (0.82-0.95) | 0.90 (0.83-0.98) |
|  | First pregnancy | 0.99 (0.86-1.14) | 0.87 (0.75-1.01) | 0.98 (0.84-1.14) |
| CVD overall | All identifiable pregnancies | 1.11 (0.96-1.29) | 0.98 (0.84-1.15) | 0.92 (0.77-1.11) |
|  | First pregnancy | 1.01 (0.74-1.37) | 0.97 (0.71-1.33) | 1.00 (0.72-1.39) |
| CVD deaths: non-myocardial infarction, non-stroke | All identifiable pregnancies | 1.07 (0.90-1.28) | 1.00 (0.83-1.20) | 0.92 (0.73-1.15) |
|  | First pregnancy | 1.04 (0.72-1.50) | 1.05 (0.71-1.53) | 1.12 (0.75-1.66) |
| Myocardial infarction | All identifiable pregnancies | 1.16 (0.83-1.63) | 0.95 (0.66-1.36) | 0.98 (0.63-1.53) |
|  | First pregnancy | 0.82 (0.34-1.98) | 0.66 (0.27-1.64) | 0.84 (0.33-2.13) |
| Stroke | All identifiable pregnancies | 1.22 (0.89-1.68) | 0.96 (0.68-1.36) | 0.89 (0.60-1.34) |
|  | First pregnancy | 1.08 (0.53-2.18) | 1.00 (0.48-2.05) | 0.79 (0.38-1.65) |
| Respiratory disease | All identifiable pregnancies | 1.40 (1.13-1.73) | 1.11 (0.88-1.39) | 0.99 (0.75-1.29) |
|  | First pregnancy | 1.48 (0.94-2.34) | 1.15 (0.72-1.84) | 1.07 (0.66-1.73) |
| Endocrine, nutritional, metabolic conditions | All identifiable pregnancies | 1.19 (0.84-1.68) | 0.95 (0.67-1.35) | 0.92 (0.64-1.32) |
|  | First pregnancy | 2.14 (1.22-3.73) | 1.44 (0.82-2.52) | 1.36 (0.75-2.46) |
| Nervous system conditions | All identifiable pregnancies | 1.11 (0.84-1.47) | 0.86 (0.63-1.16) | 1.16 (0.81-1.67) |
|  | First pregnancy | 1.06 (0.58-1.92) | 0.79 (0.43-1.47) | 0.85 (0.45-1.60) |
| **Cause-specific mortality: non-natural causes^c^** | All identifiable pregnancies | 1.22 (1.05-1.42) | 0.78 (0.66-0.92) | 0.85 (0.70-1.04) |
|  | First pregnancy | 1.39 (1.03-1.85) | 0.91 (0.67-1.23) | 1.08 (0.79-1.49) |
| Motor vehicle accident | All identifiable pregnancies | 1.07 (0.77-1.47) | 0.91 (0.64-1.30) | 0.77 (0.50-1.21) |
|  | First pregnancy | 1.09 (0.56-2.12) | 1.02 (0.51-2.07) | 1.07 (0.52-2.22) |
| Non-motor vehicle accident or violence | All identifiable pregnancies | 1.28 (1.02-1.59) | 0.76 (0.60-0.96) | 0.72 (0.54-0.96) |
|  | First pregnancy | 1.56 (1.00-2.45) | 1.01 (0.64-1.60) | 1.18 (0.74-1.89) |
| Suicide | All identifiable pregnancies | 1.33 (1.05-1.70) | 0.77 (0.59-0.99) | 1.14 (0.84-1.56) |
|  | First pregnancy | 1.50 (0.94-2.41) | 0.80 (0.49-1.30) | 1.04 (0.63-1.72) |
| All other causes | All identifiable pregnancies | 1.21 (1.07-1.37) | 0.97 (0.86-1.11) | 0.83 (0.71-0.97) |
|  | First pregnancy | 1.30 (1.02-1.67) | 1.06 (0.82-1.37) | 1.04 (0.80-1.35) |
| ^a^ Adjusted for age at the pregnancy end, calendar year of pregnancy, civil status, highest completed education, employment, and personal year-specific income, reproductive history (number of previous identifiable pregnancies, history of at least one pregnancy termination or miscarriage, hypertensive pregnancy disorders, and placental complications at previous deliveries), pre-existing chronic conditions (obesity, thyroid disorders, chronic obstructive pulmonary disease, chronic liver and kidney disease, hyperlipidaemia, hypercholesterolemia, cardiovascular, and metabolic diseases) and medication use before the index date. Analyses of pregnancies ending in childbirth were additionally adjusted for placenta-related conditions at the index pregnancy  ^b^ Data were available through 2018  ^c^ Data were available through 2017  CI, Confidence interval; CVD, cardiovascular diseases | | | | |

### **Table 5 Adjusted hazard ratios (95% CIs) for the associations between vaginal bleeding and all-cause and cause-specific mortality following the last identifiable pregnancy of a woman at age 35 years or later, Denmark, 1979-2018**

|  | **HRs (95% CI)**^a^ | | |
| --- | --- | --- | --- |
|  | **VB-affected vs VB-unaffected pregnancy** | **VB-affected pregnancy vs termination** | **VB-affected pregnancy vs miscarriage** |
| **All-cause mortality**^b^ | 1.03 (0.92-1.16) | 0.85 (0.74-0.97) | 0.80 (0.69-0.93) |
| **Cause-specific mortality: natural causes^c^** | 1.03 (0.90-1.17) | 0.81 (0.70-0.94) | 0.80 (0.68-0.95) |
| Cancer | 1.04 (0.88-1.23) | 0.85 (0.70-1.02) | 0.90 (0.73-1.11) |
| Overall CVD | 0.91 (0.61-1.36) | 0.60 (0.39-0.95) | 0.58 (0.35-0.96) |
| CVD other than myocardial infarction and stroke | 0.76 (0.43-1.35) | 0.45 (0.24-0.87) | 0.37 (0.19-0.70) |
| Myocardial infarction | 1.37 (0.59-3.19) | 0.67 (0.26-1.74) | 0.72 (0.25-2.05) |
| Stroke | 0.99 (0.48-2.04) | 1.00 (0.45-2.23) | 1.12 (0.44-2.88) |
| Respiratory disease | 0.99 (0.54-1.79) | 0.68 (0.36-1.29) | 0.68 (0.32-1.45) |
| Endocrine, nutritional, metabolic conditions | 0.69 (0.21-2.21) | 0.80 (0.27-2.39) | 0.97 (0.28-3.40) |
| Nervous system conditions | 0.97 (0.45-2.13) | 0.60 (0.27-1.35) | 0.60 (0.24-1.47) |
| **Cause-specific mortality: non-natural causes^c^** | 0.87 (0.49-1.55) | 0.56 (0.31-1.03) | 0.50 (0.24-1.04) |
| Motor vehicle accident | 0.28 (0.04-2.00) | 0.19 (0.03-1.39) | 0.34 (0.04-2.62) |
| Non-motor vehicle accidents or violence | 1.13 (0.49-2.60) | 0.62 (0.25-1.54) | 0.35 (0.14-0.88) |
| Suicide | 0.99 (0.42-2.34) | 0.68 (0.28-1.61) | 0.73 (0.25-2.12) |
| All other causes | 1.14 (0.80-1.63) | 0.98 (0.67-1.43) | 0.71 (0.46-1.09) |
| ^a^ Analyses were adjusted via IPT-weighted Cox proportional hazards regression for age at the pregnancy end, calendar year of pregnancy, reproductive history and pre-existing comorbidities (obesity, thyroid disorders, rheumatic conditions with heart involvement, chronic obstructive pulmonary disease, chronic liver and kidney disease, hyperlipidaemia, hypercholesterolemia, cardiovascular, metabolic diseases), and psychiatric conditions. Analyses of pregnancies ending in childbirth were additionally adjusted for placenta-related conditions at the index pregnancy.  ^b^ Data were available through 2018  ^c^ Data were available through 2017  CI, Confidence interval; CVD, cardiovascular diseases; IPT, inverse probability of treatment | | | |
